# Omicron detection with large language models and YouTube audio data

**DOI:** 10.1101/2022.09.13.22279673

**Authors:** James T. Anibal, Adam J. Landa, Nguyen T. T. Hang, Miranda J. Song, Alec K. Peltekian, Ashley Shin, Hannah B. Huth, Lindsey A. Hazen, Anna S. Christou, Jocelyne Rivera, Robert A. Morhard, Ulas Bagci, Ming Li, Yael Bensoussan, David A. Clifton, Bradford J. Wood

## Abstract

Publicly available audio data presents a unique opportunity for the development of digital health technologies with large language models (LLMs). In this study, YouTube was mined to collect audio data from individuals with self-declared positive COVID-19 tests as well as those with other upper respiratory infections (URI) and healthy subjects discussing a diverse range of topics. The resulting dataset was transcribed with the Whisper model and used to assess the capacity of LLMs for detecting self-reported COVID-19 cases and performing variant classification. Following prompt optimization, LLMs achieved accuracies of 0.89, 0.97, respectively, in the tasks of identifying self-reported COVID-19 cases and other respiratory illnesses. The model also obtained a mean accuracy of 0.77 at identifying the variant of self-reported COVID-19 cases using only symptoms and other health-related factors described in the YouTube videos. In comparison with past studies, which used scripted, standardized voice samples to capture biomarkers, this study focused on extracting meaningful information from public online audio data. This work introduced novel design paradigms for pandemic management tools, showing the potential of audio data in clinical and public health applications.

## 1. Introduction

Social media platforms have over 3.6 billion users globally and are projected to exceed 4.4 billion users by 2025.^1^ Every minute, approximately 500 hours of video is uploaded to YouTube alone, and much of this data is available to researchers through advanced programming interfaces provided by the company.^2,3^ This immense database contains unscripted “real-world” audio data, which may support the development of generalizable AI models. As such, the millions of hours of unscripted audio data which is publicly available via YouTube may facilitate advancements in artificial intelligence (AI) that surpass the limitations of existing methods.^4^

Multiple past studies have utilized social media data in the development of AI models for health-related tasks. A deep learning model trained on tweets was more predictive of atherosclerotic heart disease mortality than a conventional model combining demographic and health-risk factors.^5^ Various methods have extracted textual and visual features from tweets to predict mental health status.^6,7,8^ Other social media platforms have also been used as data sources for biomedical/clinical applications, though less frequently than Twitter/X. For example, analysis of YouTube home videos by non-clinical raters was able to identify autism in children. AI methods have also used YouTube data to screen for depression and OCD.^9,10,11^

Given the recent recognition of voice/speech as a promising tool for digital healthcare, work must be done to expand the use of readily available data from social media, supporting the development of new diagnostic tools for diseases with audio biomarkers. Previous efforts with limited datasets have highlighted the potential of these methods. In one example, COVID-19 breath sounds were characterized by unique time and frequency domain patterns.^12^ A CNN model trained on forced-cough recordings from a small number of patients with and without COVID-19 was able to recognize COVID-19 with high sensitivity, even in otherwise asymptomatic subjects^13^. AI technologies trained on cough sounds have also been deployed on a smartphone app for COVID-19 detection, and a binary classifier was able to differentiate COVID-19 speech from normal speech based on scripted telephone data.^14-19^ The spectral features of speech in asymptomatic patients with and without COVID-19 yielded a true positive rate of 70%, though the likelihood of generalization was low due to scripted collection and small sample size. ^20^

In a dynamic pandemic such as COVID-19, crowdsourced datasets allow for continuous sample collection over time. “Coswara” is a database containing respiratory sound samples, including cough, breath, and voices reading standardized scripts.^21^ The samples, which were recorded and uploaded by volunteers, were divided into COVID-19 (self-reported positivity) and non-COVID cohorts.^21^ Numerous researchers have used this database to train AI models for COVID-19 detection, often obtaining very high accuracies on limited binary datasets, which often excluded other respiratory illnesses^14,17,22,23,24^. Results from deep learning models trained on the “Sounds of COVID” dataset showed that voice alone led to poor performance on pre-Omicron COVID-19 data (0.61 ROC-AUC score).^4^

Most studies involving audio data have focused on the development of general AI models for COVID-19 detection without specifying the variant. Bhattacharya et al. used Coswara data to show that there were voice changes which may separate Omicron samples from healthy controls and the Delta variant, but other respiratory illnesses were not included in the study, limiting applicability. ^25^ The COVYT dataset contains COVID-related videos from YouTube and TikTok with control samples from the same speakers. However, COVYT does not include other illnesses and does not account for significant differences in content between the two cohorts (discussions of illness and random topics), which limits the potential for robust analysis.^26^

The Omicron variant of COVID-19 brought a new factor to the equation. This variant has a reduced affinity for the lower respiratory tract and, rather, affects the upper respiratory tract, particularly the vocal cords, causing symptoms of laryngitis.^27^ The changes observed in this variant, which represented a new phase in the COVID-19 pandemic, made Omicron a key area of study in the development of AI for infectious diseases with rapidly shifting symptoms and severity.

This study leveraged large language models (LLMs) and weakly annotated audio data from YouTube to detect COVID-19 cases and perform variant classification.^28^ As opposed to other diagnostic methods which rely on lab results or images, AI tools trained on audio data may be more accessible, cost-effective, and deployable in diverse settings. There are, however, numerous barriers which must first be overcome. Many prior attempts have failed due to reliance on small, binary datasets, producing overfit models which do not generalize.^29^ Existing audio models for diagnostics have also been limited due to training on short, scripted samples collected in controlled environments, losing potentially valuable information contained within freely spoken language. In this report, we introduce a hierarchical pipeline for tasks involving unscripted, “real world” audio data from YouTube.

### Contributions

1. <u>Curation of a dataset containing large amounts of real-world audio from YouTube.</u> This dataset included hundreds of hours of unscripted audio across cohorts that included self-reported COVID-19 (stratified by variant), long COVID-19, other respiratory illnesses, and multiple categories of healthy controls.
2. <u>Development of a practical pipeline using LLMs and YouTube audio data.</u> Classification tasks were designed to simulate scenarios which may arise in future pandemics, beginning with interpretable detection of disease cases (COVID-19, Long COVID, other respiratory illnesses) and culminating with classification of variants.
3. <u>Classification of COVID-19 variants with public online audio data collected in a pandemic setting</u>. In this novel application of LLMs, predictions were made purely based on symptoms and other health-related information from audio transcripts, without any reliance on specific references to the variant name or the date. Experimental results aligned with large, expensive studies involving prospectively collected data.

In contrast to conventional voice AI, this LLM-driven system was designed to leverage information contained in unscripted audio data recorded by speakers describing their experiences with illnesses. Such a system may have the potential to peruse vast amounts of audio data from future pandemic situations, facilitating the development of customized LLMs for public health and enabling a more detailed understanding of emerging infections.

## 2. Methods

This study was approved by the Institutional Review Board (IRB). The data collection, preprocessing, and assessment components of the LLM-centered pipeline are outlined in Fig. 1.

**Figure 1:**
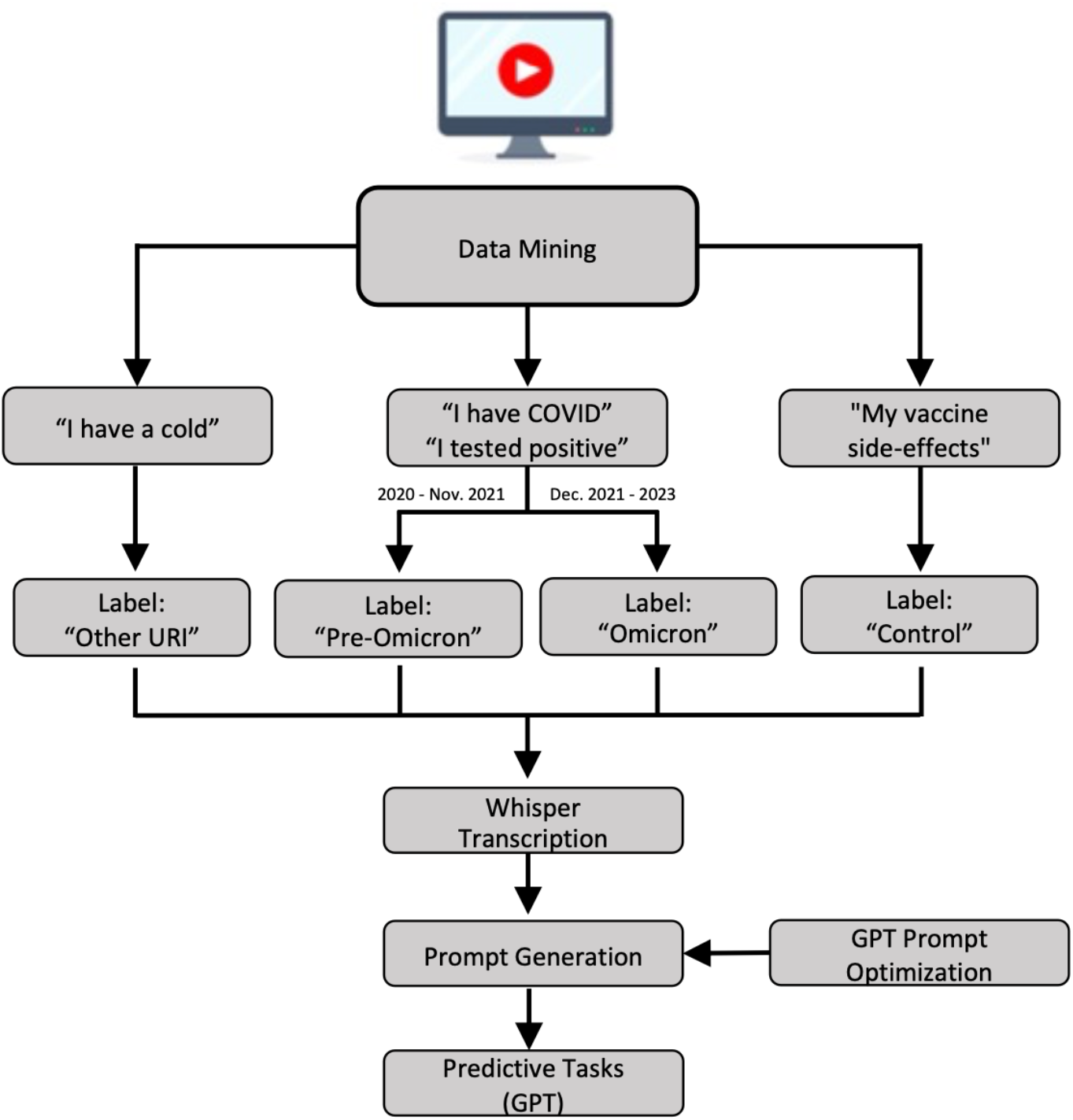
Data processing workflow used to prepare YouTube data for input into LLMs.

### 2.1 Data Collection

Videos were mined from YouTube and divided into cohorts (detailed below) based on self-declaration of health status and correlation with epidemiological data. All YouTube videos were manually verified to ensure accurate labeling based on information reported by the speaker.

**Cohort 1:** Self-reported COVID-19.

1a. Omicron variant/subvariants (presumed by dates)

1b. COVID-19 variants prior to Omicron (presumed by dates)

**Cohort 2:** Long COVID-19 (presumed by speaker declarations in the video)

**Cohort 3**: Self-reported non-COVID illnesses which frequently cause respiratory symptoms.

**Cohort 4:** 4Presumably healthy or non-acutely ill, divided into four categories:

4a. Discussions of COVID topics or experiences (e.g., testing with a self-reported negative result, vaccination side-effects), often similar in nature to the discussions of COVID-19 disease cases (1a-1b).

4b. Discussions of COVID-19 in other contexts (e.g., informatively), using similar terminology as the COVID-positive cohort.

4c. Discussions of respiratory illnesses in other contexts.

4d. Discussions of random topics such as product reviews, discussion of literature, lifestyle vlogs, and other health conditions not related to the respiratory system/viruses.

#### 2.1.1 YouTube Mining Methods

Videos which did not contain any health information were excluded from the study. For all samples in the self-reported COVID-19 cohort, the speaker explicitly confirmed a currently or previously positive COVID-19 test and discussed their experiences/symptoms related to the illness. For the cohort containing other illnesses, videos uploaded after the start of the COVID-19 pandemic were excluded if there was no self-declared diagnosis or negative test.

To facilitate variant classification, self-reported COVID-19 audio was further categorized as either “Omicron” or “non-Omicron” (Pre-Omicron). This binary labelling was done due to the relative similarity between COVID-19 variants prior to Omicron. The Omicron variant represented a shift in symptom presentation and transmissibility. Omicron was designated a “variant of concern” on November 26^th^, 2021 by the WHO, and was estimated to be the dominant variant in the U.S. by late December 2021.^30,31^ Soon after, Omicron was identified as the dominant variant globally, accounting for > 98% of sequences shared on GISAID after February 2022.^32,33^ All videos labeled “Omicron” were recorded on or after December 1, 2021 and include both the original Omicron variant and subvariants. All videos recorded on or prior to November 30, 2021, were labeled as “non-Omicron.” Table 1 lists the inclusion criteria for each cohort.

**Table 1:**
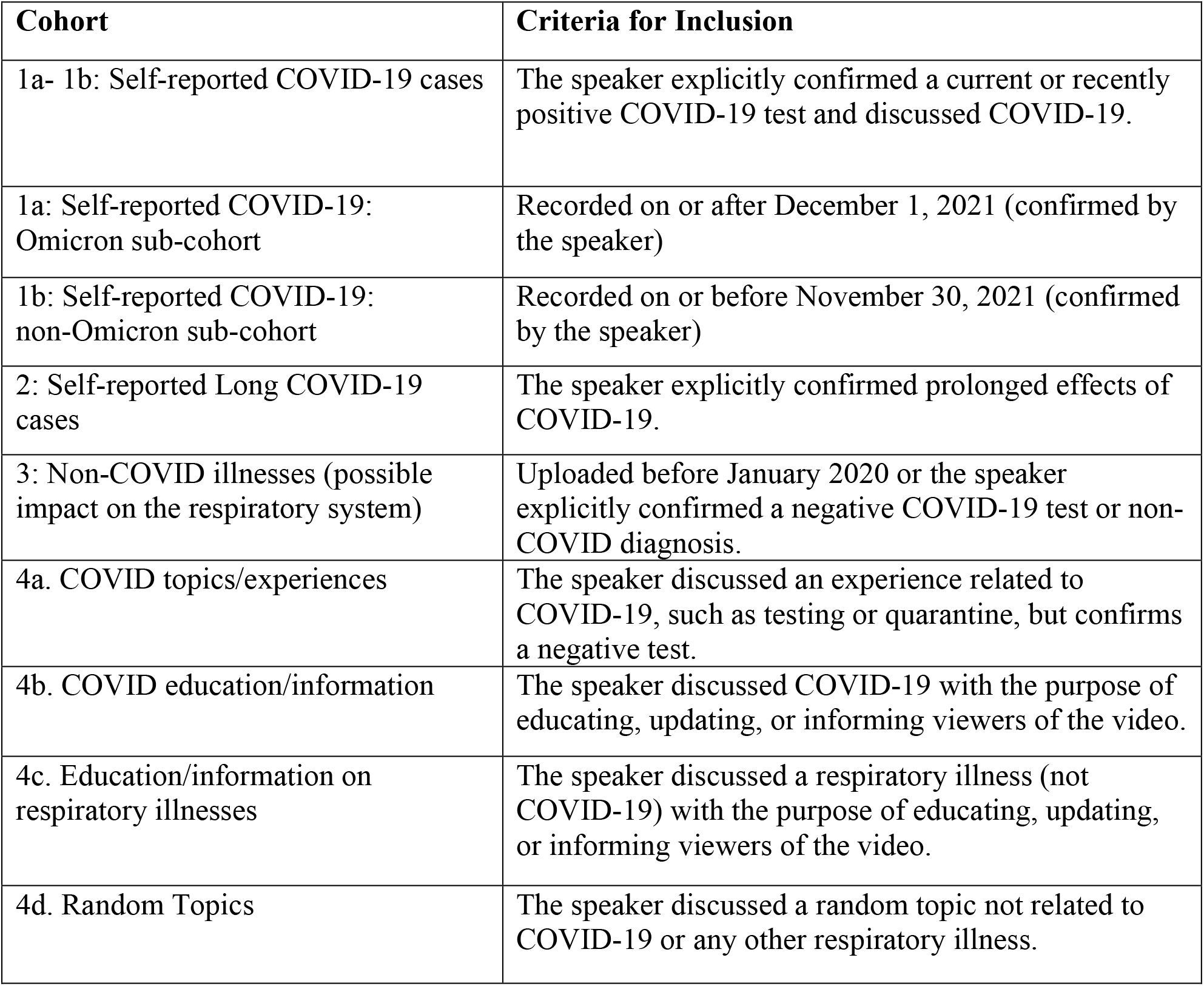
Inclusion criteria for the cohorts of YouTube audio data defined for this study.

### 2.2 Automatic Speech Recognition

Automatic speech recognition (ASR) was performed using the large pre-trained Whisper model (OpenAI), which generated a transcript for each of the YouTube videos.^34^

### 2.3 Prompt Engineering

Prompt engineering was performed to ensure optimal performance of pre-trained LLMs on transcribed information from YouTube audio data (Table 2). GPT-4 was used to refine each prompt prior to use in the LLM pipeline. ^35^

**Table 2:** GPT-4 optimized prompts used for LLM-driven detection of COVID-19 and variants.

| Category | Prompt |
| --- | --- |
| COVID-19 Identification | Examine the transcript to determine if the personal experiences described by the speaker suggest a current or previous COVID-19 infection. The following are not to be considered indicators of COVID-19:<br>1. Personal discussions about side-effects from vaccines, symptoms after vaccination, or other experiences related to vaccination. "<br>2. Personal discussions about experiences related to health but not an illness. For example, diagnostic testing with a negative result. "<br>3. Any content which does not directly pertain to the speaker discussing their own experience suffering from an illness. Topics include educational or informational content, disease pathology, symptoms, health advice or trips, recommendations from health professionals, pandemics, vaccinations, diagnostic testing, health policies, case studies, treatment options, technologies, or scientific research.<br>If the transcript mainly contains content related to any of the above (1-3), the answer should always be 'No COVID'. Summarize your conclusions in a Python dictionary format: |
|  | 'Disease status': 'COVID' (if the speaker is discussing their personal experience suffering from a COVID-19 infection), 'Long COVID' (if the speaker is discussing their personal experience suffering from prolonged effects of a COVID-19 infection), 'No COVID' (if the speaker is not discussing their personal experience suffering from COVID-19), or 'Impossible to Speculate' (if insufficient information is provided).<br>'Keywords': Rank the speaker's mentioned symptoms and health factors by their relevance to the analysis.<br>This is a simulation to evaluate AI's text interpretation abilities and not for actual medical diagnosis. |
| Variant Classification | Analyze the provided COVID-19 positive case transcript to predict the virus variant, focusing strictly on symptom-related information. Exclude direct mentions of the variant name and any date information from the analysis. The prediction should be based on symptomatology and health-related factors mentioned in the transcript.<br>Output the analysis in as a python dictionary with the following structure:<br>Variant: This should be a string value, one of 'Omicron', 'Non-Omicron', or 'Impossible to Speculate'. Choose 'Omicron' or 'Non-Omicron' based on the symptoms and other health-related factors detailed in the transcript. Use 'Impossible to Speculate' if the transcript lacks enough information for a prediction.<br>Keywords: An array of strings, listing the most relevant symptoms and health-related factors from the transcript that influenced your prediction. Arrange these keywords by their importance to the analysis.<br>This task is hypothetical and serves as an AI model exercise. It is not a medical diagnosis nor should it replace professional healthcare advice. |

#### 2.3.2 COVID-19 Detection

Prompts were designed for instructing LLMs to determine the likelihood of a COVID-19 diagnosis based on transcribed audio data. The model was told to focus on personal symptoms and other experiences related to illness, excluding educational/informative content, discussions of vaccine side-effects, or other experiences related to COVID-19/health (e.g., pre-travel COVID-19 testing with a negative result). If the LLM was unable to find sufficient evidence of COVID-19 positivity, the model was instructed to assign “Long COVID”, “No COVID”, or “Impossible to Speculate”, depending on the information contained in the transcript.

#### 2.3.3 Variant Classification

For audio samples which were predicted to be in the self-reported COVID-19 cohort, the model was further prompted to identify the specific variant. The model was instructed to ignore any mentions of specific variant names (“Delta”, “Omicron”) and any references to the date. Since the cohorts were annotated based on dates alone (i.e., audio samples from 2020 would not be in the Omicron cohort), this information could serve as a major non-clinical indicator of variant status, providing a false signal for the model and skewing the results.

For each task, the prompt was designed to ensure that the LLM would output the list of keywords used to make the final decision, facilitating enhanced understanding of the diseases. Moreover, due to the weakly annotated nature of dataset (self-reported illnesses; no confirmed test results) and the variability of information density in YouTube videos, the model was also given the option to indicate uncertainty, reducing the likelihood of meaningless random guessing. A visual overview of the data preprocessing workflow can be found in Figure 1.

### 2.4 Experimental Design

Experiments were performed to determine the potential of social media data in supporting the development of AI models for pandemic preparedness and public health tasks (in this case, COVID-19 case detection and variant classification). In these experiments, the prompts defined in section Table 2 and Whisper-generated transcripts of YouTube videos were used as inputs to the LLM. The accuracy of the LLM is reported for each cohort of audio samples (section 2.1). The LLM was also instructed to return the words/terms which were most valuable in the classification of the audio sample. Experiments in this study were performed with GPT-3.5 and GPT-4, powerful LLMs pre-trained on internet data with demonstrated complex reasoning capabilities. ^35^ To ensure reproducible outputs, the temperature parameter of GPT was set to zero.

#### 2.4.1 COVID-19 Detection

To understand the viability of audio AI models for pandemic management and other public health tasks, the LLM pipeline was first used to perform a general COVID-19 detection task with the YouTube data. The LLM was prompted to parse the provided transcript and determine if there was evidence of a past or present COVID-19 case based on health information reported by the speaker. Long COVID-19 was classified separately, as this is a distinct condition requiring separate consideration. The model was allowed to indicate uncertainty if there was insufficient information within the transcribed audio data to decide the disease status (e.g., “runny nose” was the only reported symptom). If there were multiple videos recorded by one speaker, these were grouped into “cases” (videos from the same period of time). The case was labeled as “COVID-19” if evidence of COVID-19 was found in one or more of the audio samples.

#### 2.4.2 Variant Classification

A second set of experiments was performed to explore the potential of YouTube audio data for the challenging task of COVID-19 variant classification. Here, the dataset contained only the audio transcripts which the LLM had previously predicted to be in the COVID-19 cohort (2.4.1). The LLM was prompted to use symptoms and other health data (excluding variant names and dates) to predict the variant status as either “Omicron” or “non-Omicron” (Pre-Omicron). The model could also indicate uncertainty, accounting for cases where there was sufficient evidence to indicate the presence of COVID-19 but not enough to stratify by variant. Like the COVID-19 detection task, audio samples were grouped by case (multiple videos from the same speaker and the same time period). Here, since the primary goal was to detect and characterize a variant of interest, the case was labeled as “Omicron” if the Omicron variant was detected in one or more of the audio samples.

## 3. Results

### 3.1 YouTube Audio Dataset

To the best of our knowledge, the YouTube dataset currently contains the largest amount of raw COVID-19 audio (over 100 hours) and audio from other respiratory illnesses that were described as non-COVID. This is in comparison to publicly available “COVID-19 sounds” datasets. The Coswara dataset contains samples which may be other upper respiratory infections but were not confirmed or self-declared as non-COVID.^21^ The COVYT dataset contains 65 samples, with less than 3 hours of self-reported COVID-19 audio, and has no data from other illnesses.^26^ In contrast, the YouTube dataset contains audio samples from multiple respiratory illnesses that were designated non-COVID based on self-declaration or date of posting (before Dec. 2019). These include influenza, strep throat, mononucleosis, tonsillitis, common cold, bronchitis, undiagnosed laryngitis (before 2020), and others.

The healthy control cohort of the YouTube dataset contains multiple distinct sub-populations. The “COVID experiences” cohort contains recordings of discussions about related topics such as diagnostic testing, with similar vocabulary as many of the samples in the COVID cohorts. Also included are recordings which focus on COVID-19/illnesses but in different, often more formal contexts (e.g., informative videos from clinics) and discussions of random topics recorded by presumably healthy speakers. This diverse range of controls may maximize the opportunity to assess the true potential of audio data and LLMs in the task of COVID-19 detection from social media datasets. The use of random videos alone as healthy controls may lead to artificially high-performance statistics – an AI model may easily determine these speakers do not have COVID-19 but perform poorly on healthy controls discussing a recent experience with PCR testing or side-effects from vaccination.

The YouTube dataset also presents an opportunity to explore the performance of AI models for diseases with variable severity, which is an unsolved challenge. Many of the self-reported COVID samples in the YouTube dataset were from mild or moderate cases (those with severe illness are unlikely to post a YouTube video). In many cases, conventional AI methods have performed best on data from severe cases. Nonetheless, mild illnesses are still contagious. Exposure to a misdiagnosed case of a mild illness may lead to a severe illness.

**Table 3:**
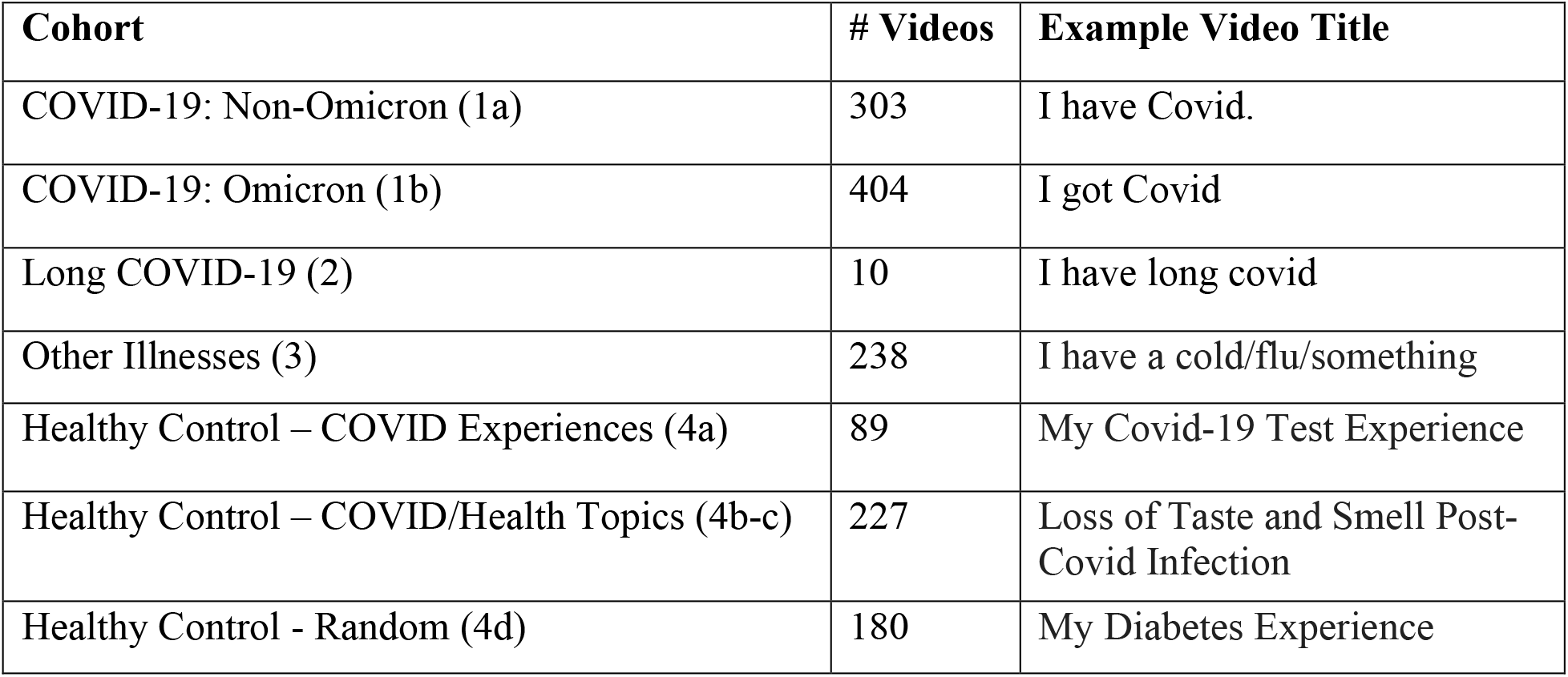
Statistics for cohorts in the YouTube dataset.

| Cohort | # Videos | Example Video Title |
| --- | --- | --- |
| COVID-19: Non-Omicron (1a) | 303 | I have Covid. |
| COVID-19: Omicron (1b) | 404 | I got Covid |
| Long COVID-19 (2) | 10 | I have long covid |
| Other Illnesses (3) | 238 | I have a cold/flu/something |
| Healthy Control – COVID Experiences (4a) | 89 | My Covid-19 Test Experience |
| Healthy Control – COVID/Health Topics (4b-c) | 227 | Loss of Taste and Smell Post-Covid Infection |
| Healthy Control - Random (4d) | 180 | My Diabetes Experience |

### 3.2 LLM Results

LLMs were used to perform hierarchical classification tasks with transcribed YouTube videos. The model was first instructed to assess if the speaker is likely discussing a past or present experience with COVID-19. For all transcripts with this designation (COVID-19 positivity), the LLM then predicted the variant based on described symptoms and health-related information.

#### 3.2.1 COVID-19 Detection

Model performance is reported for the self-reported COVID cohorts as well as subgroups of COVID-negative controls. This initial task involved predicting the likelihood of a COVID-19 infection using information from the transcript, focusing on symptoms, interventions, and other related factors (e.g., quarantine periods, specific instructions given by HCWs).

**Table 4:** LLM performance across sub-cohorts within the YouTube dataset.

| Cohort | Desired Outcome | Accuracy |
| --- | --- | --- |
| COVID-19 Positive | COVID-19 | 0.89 |
| Long COVID-19 | Long COVID-19 | 0.90 |
| Other Resp. Illnesses | No COVID-19 | 0.97 |
| COVID Experiences/Topics | No COVID-19 | 0.97 |
| COVID/Resp. Information | No COVID-19 | 0.96 |
| Random Topics | No COVID-19 | 0.99 |

Results show that GPT models can identify COVID-19 infections and Long COVID-19 in speakers discussing their experiences with an illness. If properly instructed, LLMs were not confused by topics such as COVID-19 testing with a negative result, vaccination side-effects, or other types of respiratory illnesses. The error rate was very low across all cohorts, and uncertainty rates were less than 1%.

#### 3.2.2 Variant Classifications

The LLM was further tasked with predicting the variant (Omicron or Non-Omicron) based on only health-related information in the transcript, excluding specific variant names and references to the date. Due to the challenging nature of this task, the model was permitted to respond with uncertainty if the transcript did not contain sufficient information to reasonably predict the variant. Uncertain cases were not considered in the calculation of accuracy statistics, as these would not be included in downstream applications of the pipeline. To best simulate real-world use, false positives from the COVID-19 detection task were included in the variant classification cohort. However, these samples were given the target label of “impossible to speculate”, since the specific symptomology information needed to predict the variant should be non-existent if the speaker is not discussing their experience with COVID-19. GPT was able to generate a prediction for over 70% of the dataset and achieved an accuracy score of above 0.75 for both the Omicron and non-Omicron cohorts. The uncertainty rate in this case was expected: YouTube videos are extremely variable in terms of information density and overall coherence. For all false positive cases, the model indicated that there was insufficient evidence for a prediction.

**Table 5:** LLM performance in variant classification (accuracy at achieving desired outcome).

| Cohort | Desired Outcome | Accuracy | Uncertainty |
| --- | --- | --- | --- |
| Omicron | Omicron Variant | 0.76 | 0.29 |
| Non-Omicron | Non-Omicron Variant | 0.78 | 0.22 |

#### 3.2.3 Interpretation

In addition to the disease state predictions, the LLM was instructed to output the key words/phrases which guided the decision-making process. These outputs are visualized as word clouds in Fig. 2-3, showing the information involved in LLM-driven identification of COVID cases, Long COVID cases, and variants. Stopwords (“a”, “an”, “the”, etc.) and other common terms (e.g. “symptom”, “positive”, “test”) were removed for clarity.

**Figure 2:**
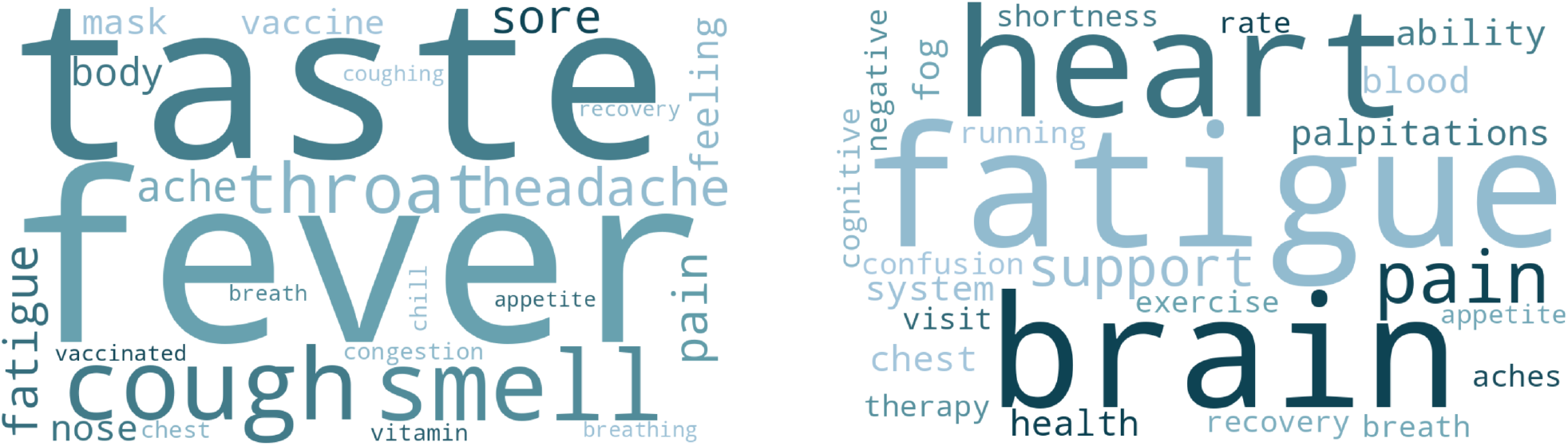
Interpretation of results from LLM-driven COVID and Long COVID detection. **Left:** keywords from non-omicron predictions. **Right:** keywords from Omicron predictions.

Fig. 2 presents the visualization of relevant terms in the task of detecting general COVID-19 cases and Long COVID-19, showing the transition from common symptoms such as fever or loss of taste (Fig. 2 - left) to cognitive and/or cardiovascular effects (Fig. 2 - right)., consistent with existing scientific literature.^36^

Figure 3 displays the interpretation of results for the variant classification task. In Fig. 3 (left), results show that LLM decision-making emphasized symptoms such as the loss of taste and smell, which aligns with large-scale studies (e.g., Menna et al.). ^37^ Compared to the non-Omicron cases (Fig. 3 - left), the keywords from Omicron predictions shown in Fig. 3 (right) significantly emphasized upper respiratory symptoms – “sore throat” and “cough” – which were found to be associated with the Omicron variant. Other keywords such as “mild” and “vaccinated” (indicating a breakthrough case) also match with past studies. ^38^

**Figure 3:**
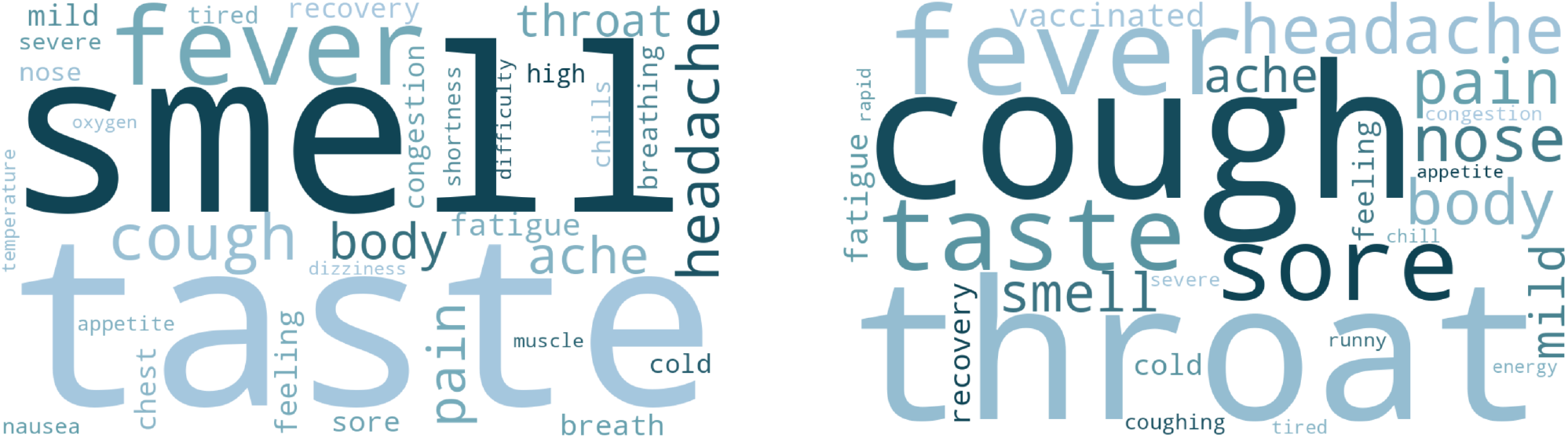
Interpretation of results from LLM-driven variant classification. **Left:** keywords from non-Omicron prediction. **Right:** keywords from Omicron prediction.

## 4. Discussion

This report shows that audio data from online sources, including noisy social media data, has potential value in pandemic settings or other public health applications when paired with powerful LLMs like GPT. LLMs may be able to perform pandemic-related tasks using audio data from social media sources such as YouTube. The LLM used in this study obtained interpretable results on detection of self-reported COVID-19 cases, including Long COVID-19, and a variant classification task. In the future, this type of system may be used to identify cases which do not fit into any existing category of variant or illness (“emerging infection identification”), or better understand vaguely defined conditions such as Long COVID-19, providing timely information to healthcare workers or public health officials responsible for pandemic management. This work may also have relevance in low-resource settings where extensive prospective data collection is infeasible and models from high-income areas are often trained on narrow datasets. The vast body of knowledge contained in LLMs may facilitate more nuanced insights with geographical and cultural context.

### 4.1 Real-World Deployment

Potential future directions for this work include the use of LLMs to facilitate epidemiological tasks, variant detection, dataset expansion through task-specific mining of social media, and encoding of weakly supervised data to facilitate fine-tuning on more reliable prospective datasets. For example, the pipeline described in this report could be used to detect and annotate vast amounts of anonymized public data from multiple online sources, expanding on the dataset used here, which is extremely limited compared to the data available across YouTube, Twitter/X, Meta, and other social media platforms used worldwide. Moreover, this study underscored the inherent multimodality of audio data, potentially spanning voice, speech, and language. Highly scalable prospective data collection studies could be run to enhance the multimodality of crowdsourced voice data, which would then be analyzed by custom LLMs as described above. This concept has been actualized in our ongoing project, which can be accessed at hearai.org and is being focused towards low-resource healthcare settings.

### 4.2 Limitations

The impact of this work currently remains highly speculative due to possible biases in the dataset and weak annotations based on self-reporting by speakers in YouTube videos. The dataset was developed only based on the availability of YouTube videos meeting the inclusion criteria (Table 1); due to the uncontrolled nature of social media data, biases are likely present (e.g., age group) in this initial proof-of-concept study. There were also additional limitations of the study due to the data collection and annotation strategies required to build the YouTube dataset. Variant status could not be confirmed serologically but was inferred based on the date of recording. The “Long COVID” cohort was limited in size and larger studies are needed to determine the role of LLMs in understanding poorly defined health conditions. The cohort of non-COVID respiratory diseases was also not serologically confirmed and may have included false negative COVID-19 cases within the subset of videos recorded after the start of the COVID-19 pandemic. Finally, in future work, experiments should be performed involving knowledge cut-offs (by date) for the LLM to assess the performance of the model at different stages of a pandemic or other health crises.

## 5. Conclusion

Ultimately, digital health is understudied in the context of multimodal audio data, social media, and LLMs. However, the wide availability of such data and the unprecedented capabilities of models like GPT raises important new questions for data rights, privacy and security, regulation, ethics, and real-world validation. As audio/video communications rapidly increase in popularity, unscripted audio data from social media may be more available and inherently more diverse than conventional modalities, leading to robust insights with clinical relevance. Even without gold standard annotations, the results achieved by this early effort at COVID-19 detection and variant assessment merit further evaluation in specific public health settings with unmet needs, particularly in low-resource settings with limited capacity for prospective data collection. Despite limitations, this work highlights the potential of unscripted audio data to enable LLM-driven automation of complex tasks involving public health and pandemic-related challenges, well beyond COVID-19.

## Data Availability

Data will be made available for academic research upon reasonable requests directed to the corresponding email.

